# A Training-of-Trainers Program for Nurses in Tanzania: ICU Standards of Care, Documentation, and Communication

**DOI:** 10.1101/2024.04.14.24305628

**Authors:** Grace Kistner, Shannon Macfarlan

## Abstract

We present one method of a Training-of-Trainers (ToT) program supported by a partnership between a low-middle income country (LMIC) hospital and a high income country (HIC) organization through lectures, group discussions, assessments, and bedside coaching over a period of two weeks. The goal was to build capacity in ICU nurses by teaching standards of care and documentation, improving interdisciplinary communication, and scaling up participants’ knowledge and comfort levels in providing quality care. Nurse educators from the Alliance for Global Clinical Training (Alliance) designed the program and conducted the critical care nursing curriculum. Topics were selected by Muhimbili National Hospital (MNH) nursing peers who also provided facility information. Understanding what resources and infrastructure are routinely available is essential to applying concepts to practice. The MNH administrative team and nursing education liaison sought out the best suited participants. Identifying talent in participants for ToT programs is crucial to their success. Nurse participants were chosen as effective representatives of their individual units to be future agents of change. Participants described what they most wanted from the program, including: increased knowledge in documentation, communication, and overall critical care specialized training. Pre- and post-knowledge assessments tested critical care knowledge. The nursing process ADPIE (Assessment, Diagnosis, Problem, Intervention, Evaluation) and SBAR communication (Situation, Background, Assessment, Recommendation) were provided tools for standard operating procedures which enhance interdisciplinary management of care. Clear and consistent documentation with ADPIE requires clinical assessment and evidence-based diagnoses. Standardized communication with SBAR provides an organized framework to professionally relay critical information and provide recommendations. All materials were provided in an open-access format for the program to be easily replicated by the participants. A long-term goal of this training was to assess impact and sustainability.

## Literature Review

Continuing education and training opportunities for practicing nurses are essential to professional advancement. Successful endeavors can be linked to the United Nations Sustainable Development Goals (SDGs) principles of health capacity training (SDG 3.d), access to education and relevant skill (SDG 4.4), training for decent work (SDG 8.6), innovation of technology (SDG 9.b), equal representation of LMIC voices (SDG 10.6), and partnerships (SDG 17.16) ^1^. There is an identified need and desire for transformative training to upskill, build capacity, and retain the nursing workforce in LMICs ^2^. It is well reported that nursing care has a direct relationship with quality of care. High quality care is integral to Universal Health Coverage that the world strives for, and highly skilled nurses are on the frontlines providing it ^3^. Global data on the number of training programs for nurses is unknown, however there are fewer opportunities in LMICs compared to HICs ^4^. To address some of these challenges, clinicians from HICs have formed partnerships with hospitals and academic institutions in LMICs to facilitate education and training opportunities ^5^. Partnerships between hospitals in LMICs with those in HICs provide an opportunity to communicate best practices in a broad range of specialties.

## Background

One such partnership exists between the Alliance and MNH, affiliated with Muhimbili University of Health and Allied Science (MUHAS) in Dar Es Salaam, Tanzania. The Alliance is a non-profit corporation which links HIC educators with Departments of Surgery and Nursing in LMICs to improve surgical care and education ^6^. Their collaborative educational relationship with MUHAS / MNH was established in 2012. MNH is positioned to spearhead service improvement as a 1,500 bed tertiary referral center which serves as the teaching hospital for MUHAS. They average 2,000 daily outpatient consultations with 2,800 professionals including physicians, registered nurses, pharmacy and supporting staff ^7^. While the global healthcare workforce shortage continues to strain systems, the recent “brain drain” phenomenon has seen up to half of newly trained Tanzanian nurses migrate ^8^. Tanzania is one of five Sub-Saharan African countries which are projected to have the highest population growth by 2050 ^9^.

In 2017, the MNH Department of Nursing requested educational assistance, and Alliance nurse educators developed a two-day course in pre- and post-operative nursing care. That course was well received, but there was no formal evaluation. Comprehensive evaluations that cover proficiency as well as confidence are under-studied ^10^. In 2019, a second request for educational assistance was made, this time in critical care. In this manuscript, we report results from a post-graduate nursing education certificate program. This ICU Standards of Care, Documentation, and Communication ToT program touched upon common themes in interdependent education ^4^. The overall goal of the partnership was to promote critical thinking, build capacity care, and encourage interdisciplinary communication focusing on care, documentation, and communication standards - an imperative component of successful ToT programs ^5^. Initiatives in the region support open-access resource availability for continuing professional development ^11^ and studies show proactive technical assistance enhances the sustainability of ToT programs ^10^. Programs taught as a result of partnerships like the Alliance and MNH can help improve the knowledge, skills, and confidence of participants.

## Method: Program Development

The planning for this program involved four steps carried out over a period of six months. First, proposed course topics were requested by MNH nursing colleagues. In response, inquiries were made regarding demographics within the ICU at MNH including common disease profiles in the patient population. We also inquired about capacity for diagnostic testing, types of monitoring equipment available, and medications frequently used. Second, the topics were collaboratively narrowed in order to tailor the course content to be relevant to this ICU. Alliance nurse educators prepared the course materials using evidence-based nursing guidelines and up-to-date best practices. The curriculum was organized in an attempt to balance old and new material for the participants who had varying levels of nursing experience but were all working in a critical care setting. The focus included critical care components in cardiovascular, neurologic, respiratory, and metabolic systems; as well as ICU standards of care, prophylaxis bundles and end of life care. Third, the MNH liaison chose fifteen staff nurse participants (out of 800 nurses at MNH) who had leadership qualities allowing them to effectively replicate the program to train their peers. Finally, Alliance nurses created sample program agendas in order for MNH colleagues to choose a format that best met their needs. Morning didactic lecture sessions covered pathophysiology and concepts of nursing standards. Afternoon bedside coaching in the ICU wards allowed for practical application of the concepts. The topics were grouped by body system and divided into six sessions over a two-week period (Table 1).

**Table 1.**
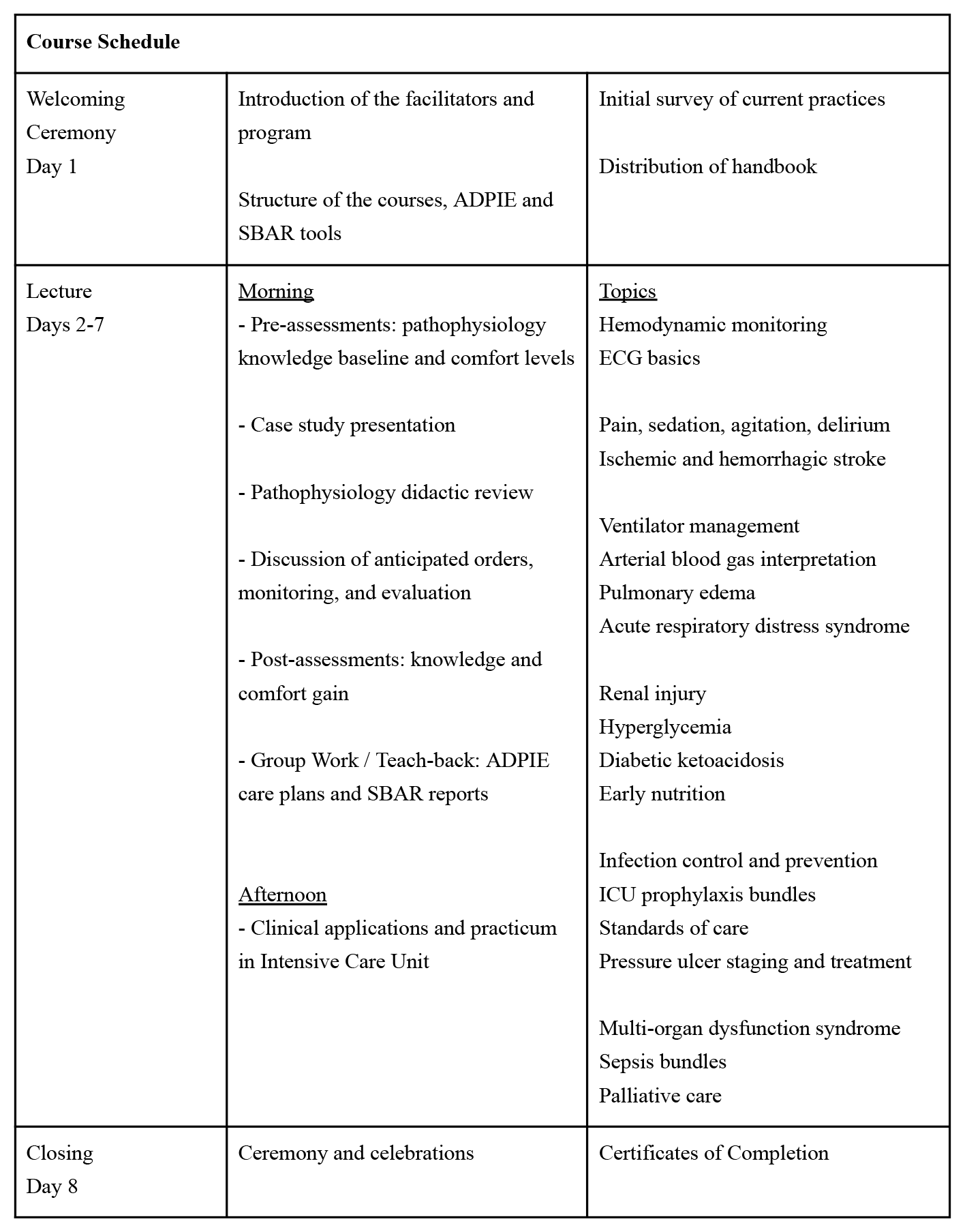
Course Schedule.

## Method: Course Description

The course materials included PowerPoint presentations on pathophysiology topics, case studies, self-assessments, and review questions. The content was designed to focus on ICU standards of care, documentation, and communication. Worksheets emphasized how to create care plans using the nursing process ADPIE ^12^. The participants had knowledge of the nursing theory but had not been incorporating it formally in their patient flowcharts. Course references included links to the North American Nursing Diagnosis Association and samples of the tool SBAR ^13^. SBAR has been widely adopted in the healthcare field to efficiently convey critical information. The first day of the course was dedicated to this communication and documentation standard framework. It focused on utilizing these tools to enhance patient-specific care and how they improve patient outcomes.

A case study was presented at the beginning of the course and each day new information was added which was relevant to the lecture topic. The participants completed pre-assessments on their knowledge of pathophysiology. A baseline understanding was established and reviewed where necessary during didactic sessions. They were then surveyed on their comfort level in caring for a critically ill patient similar to the one in the case study. The participants were posed with a series of questions on their knowledge of evidence-based information for current best practices. They were challenged to think critically about orders that might be anticipated, what focused assessments were imperative and what necessary monitoring would help determine effective treatment. Post-assessments provided data on the amount of knowledge and comfort gained after each lecture. All assessments and surveys were anonymous and no specific participant data was identified. Participants worked in groups to discuss and present teach-back of the materials to reinforce learning and practice skills teaching to peers. During the bedside portion of the training, patients were selected based on diagnoses corresponding to lecture topics. Participants assessed the patient and then return-demonstrated clinical skills using available equipment. They made nursing management recommendations incorporating ADPIE and practiced handoffs with SBAR communication.

## Findings and Discussion

An initial pre-program survey (Figure 1) was completed that aided understanding of prior documentation and communication practices as well as cultural implications in nursing education, practice, and environment. The majority of participants did not identify minimum degree requirements for ICU nursing or specific certifications required for their wards. They noted there is not a formal orientation nor regular training in their place of practice. Conversations with the participants revealed that preceptorship and of new staff sometimes lasts no longer than one month. Intern nurses were assigned patients with advanced critical illnesses on their first day.

**Figure 1.**
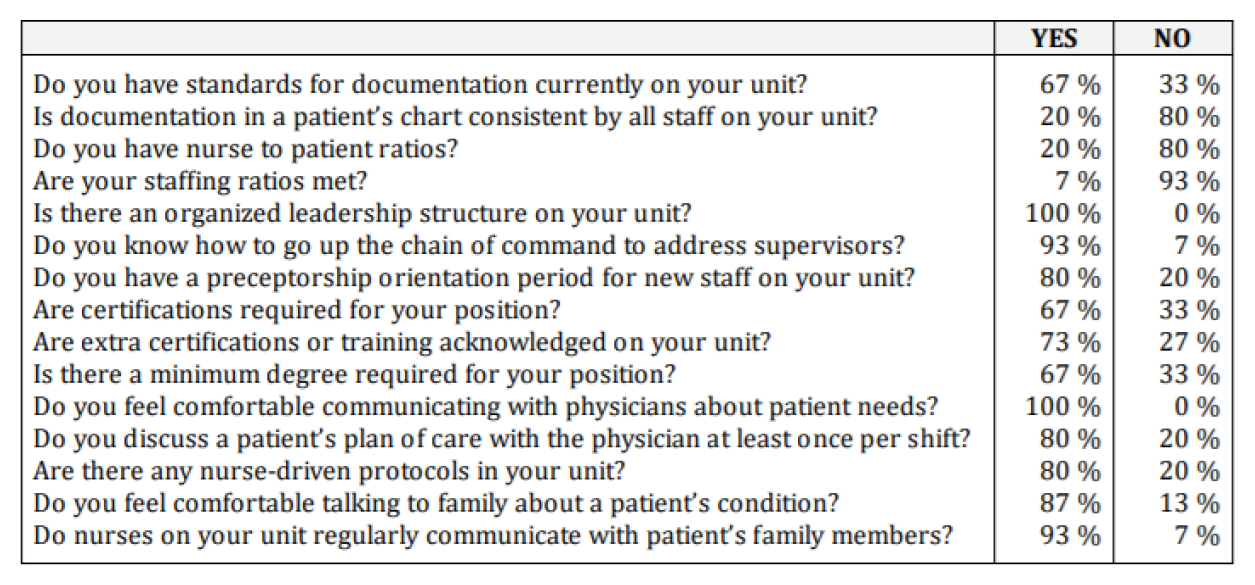
Initial Pre-Program Survey.

This information demonstrated the need for additional training programs. The participants expressed desire for increased frequency and more specialty training when asked to identify opportunities for change in their practice (Figure 2) as well as what they hoped to learn from this particular program (Figure 3). This represented a match of training needs to the content of our training program.

**Figure 2.**
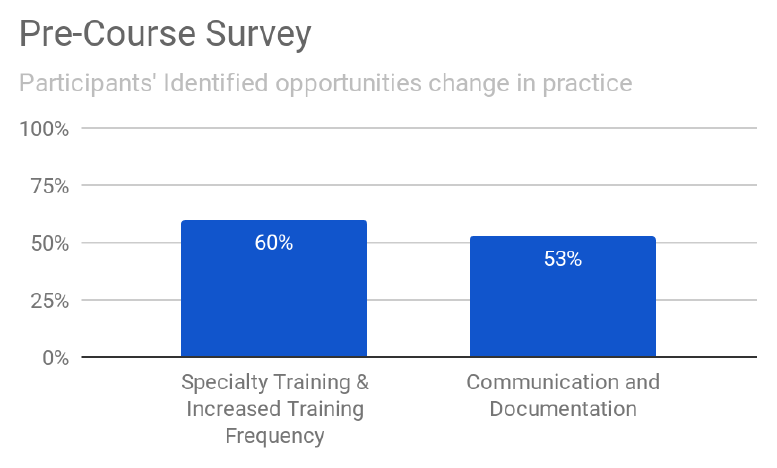
Pre-Course Survey: Identified Opportunities for Change.

**Figure 3.**
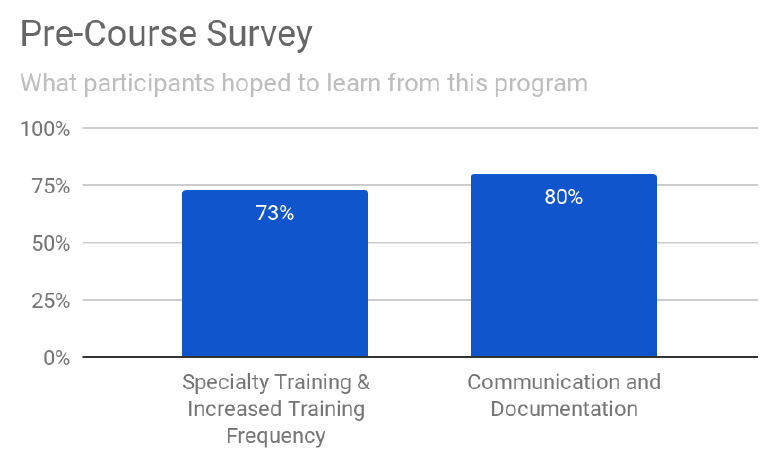
Pre-Course Survey: What Participants Hoped to Learn.

Most participants acknowledged that documentation in their wards was inconsistent as evidenced by incomplete charting or lack of documentation altogether. Although survey results indicated they felt comfortable communicating with physicians about patient care, lecture discussions revealed that physicians are not regularly available at the bedside or by phone. Furthermore, physicians did not incorporate nurses in daily rounds as a standard practice. The participants’ comfort levels utilizing the ADPIE and SBAR tools were measured (Figure 4), as well as their comprehension of applying the concepts to each clinical topic (Figures 5, 6). The findings showed the participants’ increased comfort levels with these tools taught in the course. Over 80% of the participants showed full comprehension by applying these concepts. Opportunities for change identified by the participants included making sure every patient has a care plan, writing a nursing diagnosis based on the assessment every shift, and knowing how to use SBAR communication.

**Figure 4.**
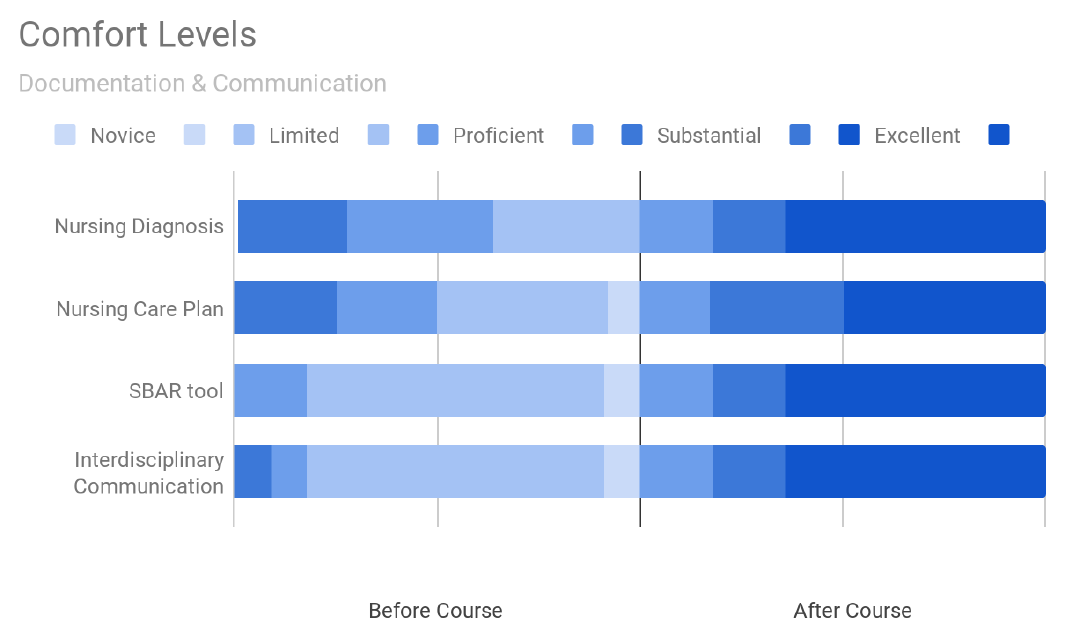
Comfort Levels: Documentation and Communication.

**Figure 5.**
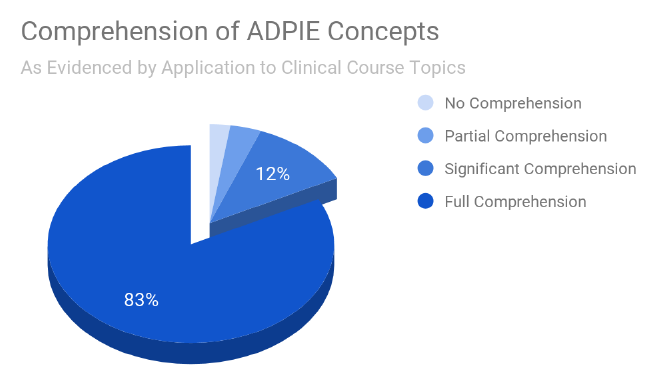
Comprehension of ADPIE.

**Figure 6.**
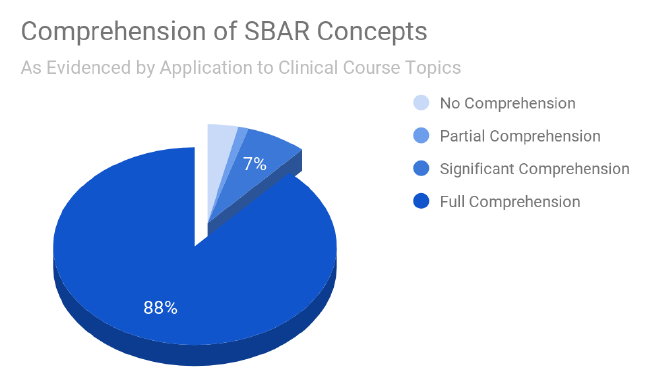
Comprehension of SBAR.

We found that the training elevated participants’ confidence levels in clinical judgement and improved standards of documentation and communication. Over 80% of the participants demonstrated full comprehension of the ADPIE nursing process for documentation and SBAR communication tools. Before the program, the majority of the participants rated their clinical knowledge and comfort levels as “Limited”. Afterwards, the majority rated their clinical knowledge and comfort levels as “Excellent”.

A review of the data shows that, overall, the identified gaps in both knowledge and skill were properly aligned to the program content. The assessment test scores (see Appendices) indicated a significant increase in comprehension as a direct result of the didactic lectures. The reported comfort levels showed the participants’ increased confidence to apply critical care nursing principles (Figure 7). The topics most notable for amelioration over the two weeks included: ECG rhythm identification and interpretation, brainstem reflex assessment, ventilator management of acute respiratory distress syndrome (ARDS), identification and management of renal injuries, implementation and contraindications of early nutrition therapy, glycemic control, staging of pressure ulcers, standards of care and documentation of invasive lines and drains, early sepsis identification and management, and palliative care.

**Figure 7.**
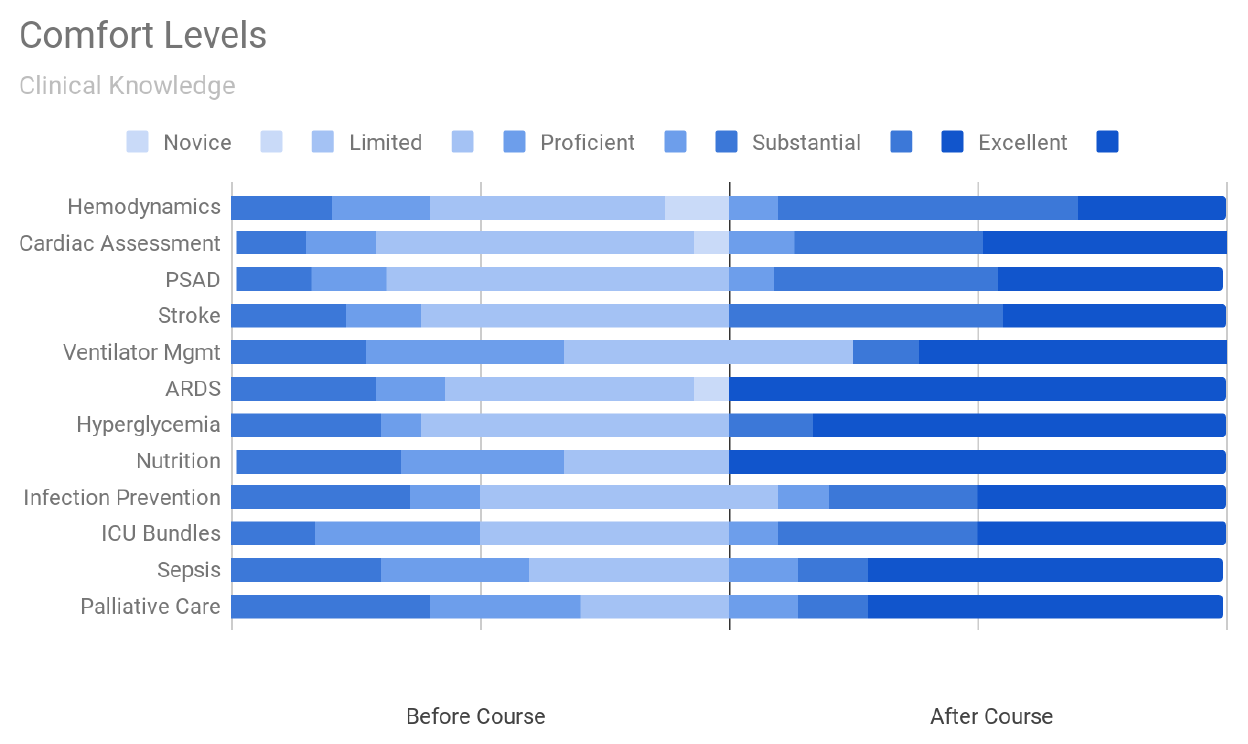
Comfort Levels: Clinical Knowledge.

The assessment results showed a 23.67% increase in the average overall score across all topics. The most gain was 38% in Metabolic Disorders and Nutrition topics. Almost one third (31.33%) of participants were able to answer all post-assessment questions correctly. Remarkably, over half (52%) of participants correctly answered all Sepsis and Palliative Care topics. Participants had never received training in palliative care before and told us that Do-Not-Resuscitate orders do not exist in their unit. Across topics, average post-assessment scores were 82% or higher.

Specific opportunities for change identified by the participants after the program included improving their skills and knowledge of cardiac monitoring, ECG interpretation, and how to intervene during hemodynamic changes. They also identified improving nursing care and assessments of stroke patients, pain management and advocacy, management of ICU delirium, scoring the patient’s Glasgow Coma Scale, and differentiating between ischemic and hemorrhagic stroke. In addition, airway and ventilator management, recognizing respiratory failure, knowledge of ARDS and interpretation of arterial blood gas (ABG) were highlighted. They also seek to implement changes in monitoring strict intake and output, identifying unique intravenous fluid needs, advocating for early nutrition, managing metabolic and renal disease, and performing regular blood sugar checks on patients. Opportunities for change related to ICU standards included not reusing single-use items, applying ICU bundles to patient care, understanding precautions for controlling the spread of infection and isolating cohorts, changing patient position every two hours, and assessing and documenting wounds. Furthermore, they noted sepsis risk factors and clinical management in accordance with evidence-based bundles. They also intended to involve the family in the patient’s care and encourage relatives to provide psychological support for their sick relatives. Finally, they indicated they would be change agents in their practice as good role models and advocates for their peers. This extensive list of identified opportunities for change demonstrated the need for specialty training to reinforce standards of ICU care. It highlights the relevance of our program to what the participants expressed desire to learn during this program.

The participants stated that as a direct result of this program, they would make changes to improve their practice to help decrease over-stay in the ICU with proper management of care. Interventions they intended to implement included moving severely ill patients near the nurse’s station for close assessment and to teach colleagues about standardized care plans. They also intended to implement monitoring of the cardiovascular system closely, teaching the other staff on the unit to interpret ECG results, and how to intervene for observed abnormalities. They will implement adequate pain control techniques for patients, proper sedation evaluation, and daily assessments to identify early neurological changes. Another key area involved ventilator management and following infection control bundles, protecting the airway, and early weaning. Proper fluid balance, early identification and management of renal disorders, and early nutrition will be implemented. A specific practice improvement identified as a direct result of the class was to avoid fluid overload in patients with acute kidney injury. They seek to adhere to standard precautions for infection prevention, advocating for compliance, performing aseptic dressing changes, choosing proper wound medication, improving handwashing in the ICU, preventing decubitus ulcers, and ensuring personal protective equipment availability. Specific practice improvements as a result of the class included improving infection prevention standards, assessing a patient’s skin and invasive lines or tubes, properly assessing and identifying early signs of infection. One participant shared “I hope [the instructors] come back next time, best presenters and materials!”

Finally, the instructors and program were rated. The participants’ feedback indicated the instructors had knowledge and skills of learning and teaching methodology, were able to provide easy examples, able to clarify and explain clearly answers to any questions asked, able to demonstrate teaching skills to the class, provided useful knowledge and skills to improve bedside practice, and were easy to understand (Figure 8). The participants gave an overall rating to the program, as well as their ease of understanding it, if it successfully provided a balance of new and old material, if it promoted excellence in nursing practice, and if the participants would recommend this program to their colleagues (Figure 9).

**Figure 8.**
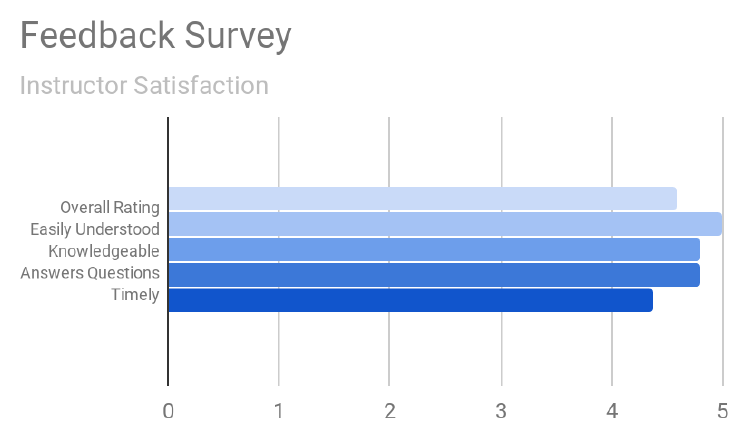
Feedback Survey: Instructors.

**Figure 9.**
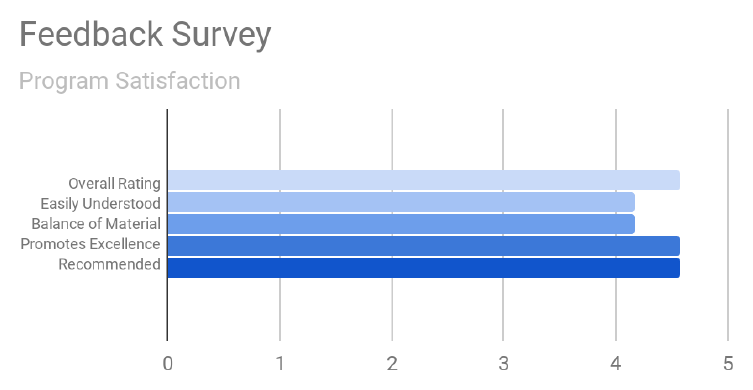
Feedback Survey: Program.

## Implications of Success

This limited study describes a proof of concept for partnerships like this to provide a focused opportunity to improve the knowledge, skills and confidence of critical care nurses in LMICs. The program was successfully designed in a collaborative effort between the Alliance and MNH / MUHAS which concentrated on pre- and post-surveys, assessments, and feedback. The goal to cover a variety of specialty topics in critical care nursing was achieved. Interpreting the results of the program data revealed that knowledge and competency gaps were significantly reduced. Survey data confirmed that the gain in knowledge was desired and the course content was beneficial and relevant. Even though the nurse participants had already been practicing, this program was successful in building capacity. Directly following the program, preliminary results showed higher levels of comfort in implementing advanced nursing care; thereafter they reported referencing the program when they were asked if they had specialty training. Tools were provided that enhanced standards of care, reinforcing documentation with ADPIE and empowering nurse involvement in interdisciplinary management using SBAR communication. Comprehensive manuals and resources were provided in an electronic, open-access format for ease of continued use and replication of the ToT.

> **MNH blog**: “Nurses in the specialized intensive care unit (ICU) of Muhimbili National Hospital (MNH), Swords and Mloganzila, have been specially trained to build capacity to care for patients and to keep records of patients admitted in those wards. The two-week training conducted in partnership with MNH and the United States’ [Alliance for Global Clinical Training], concluded today with 15 nurses having been awarded certificates of [completion] … 15 nurses were selected from the 800 nurses for training and they would train other nurses. [Mr. Munguatosha said] ‘*I expect to see a positive change in your performance as I believe you are going to be your fellow teachers in your workplace and this will improve your performance*.’ And the trainer from the [Alliance], Grace Shaw [Kistner], thanked MNH’s leadership for their cooperation throughout the training.” ^14^

During the closing ceremony, certificates of completion were awarded to the participants. They stated that the program promoted excellence in practice to provide quality care and one participant added, “My patient will be managed as an individual” as a result of the program. The main liaison from MNH nursing indicated with satisfaction that the program aligned with her continued efforts of implementing ICU standards of care. The MNH nursing administration echoed the Alliance volunteer nurse instructors’ expectations for improvement in clinical practice and delivering this educational program to their peers. A translated excerpt from the MNH website highlighted the implications that the training would have on nursing care.

## Limitations and Opportunities

Limitations were realized in this first ToT program developed by Alliance nurses. Lessons learned were heavily weighted in the need to have a better baseline understanding of the participants knowledge, equipment available, and established policies. Pre-assessment surveys should be completed earlier in the planning process to identify knowledge gaps and customize course content, and a bilateral effort is required to facilitate a needs assessment. Cultural and organizational challenges were encountered revolving around healthcare in Tanzania and the adoption of new concepts taught in the course. The results of our initial survey indicated that there was no established nurse to patient ratio. An increased number in patient workloads may not allow adequate time for consistently implementing techniques from the course. This can negatively impact care, documentation, and communication despite increases in knowledge or skills. All of these constraints should be considered in subsequent pre-trip needs assessments, factoring in what U.S. or other HICs standards might need to be adapted to meet local policy and procedure guidelines, healthcare laws, and culture. Despite these challenges, nursing leaders welcome assistance in learning best practices for optimum care and outcomes.

Opportunities for improvement in a future program include narrowing content to more specific educational needs, tracking patient data, and direct observation of participants training their peers. Future topics for consideration discussed while in-country included: narrowing pathophysiology topics for review, expanding protocols, specialty training, and collaborative reviewing of policies. Tracking data before and after these programs can help determine if there is a direct impact of the upskill on quality of care. Data to track might include mortality and morbidity rates, chart audits, and communication tools. More time could be allocated for a formal audit of the participants teaching their peers. Observing and evaluating training sessions would be an indicator for continued ToT program success. Long term performance review and continued nurturing is needed to assess sustainability of ToT programs. The follow up actions requested of MNH included: a) complete a post-trip feedback questionnaire (Figure 10), b) identify participants who have taught a ToT to their peers using the course materials provided and c) report on the outcomes of that program.

**Figure 10.**
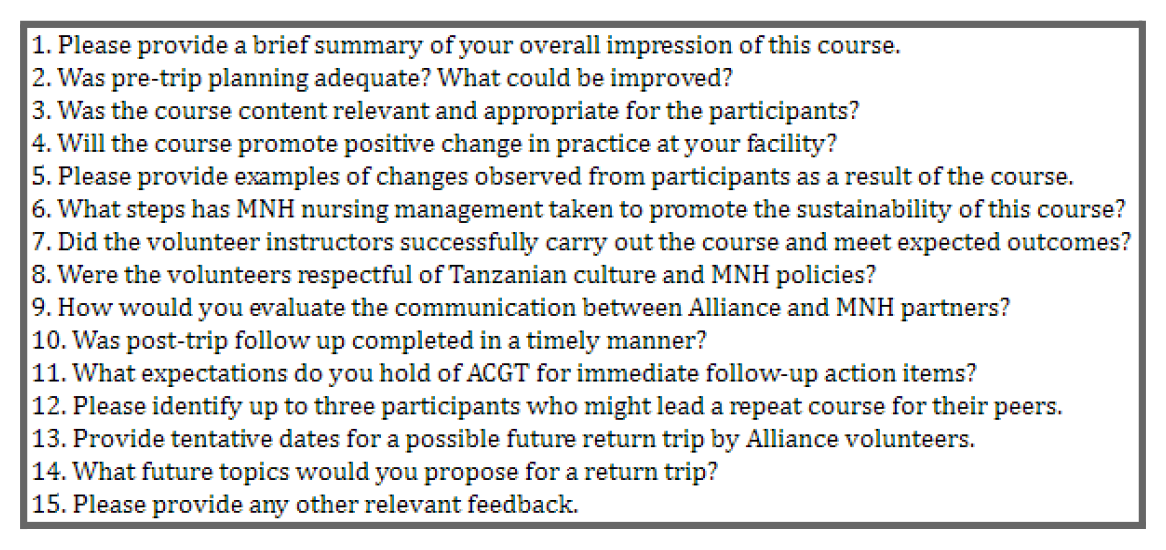
Post-Trip Feedback Questionnaire.

Unfortunately, due to the SARS-CoV-2 outbreak, international travel bans hindered plans for a timely follow up of this training in-person. Online materials and support continued to be available throughout the pandemic.

## Data Availability

All data produced in the present work are contained in the manuscript and its appendix

## Acknowledgements

We would like to thank the board officers of the Alliance, who recognized that their physician surgical volunteers had been well-received at MUHAS/MNH and were having an important influence on enhancing the surgical skills of their counterparts. As a result, surgical leadership at MUHAS/MNH agreed they would support assistance to contribute to nursing care improvements. Ultimately, this ignited the Alliance Nursing Education Initiative. We would also like to thank leadership from the MNH nursing administration office under the guidance from Zuhura Mawona (Ag. Director of Nursing) and Yassin Munguatosha (Head of Nursing Department). A special thanks goes to the primary liaison for this program Trustworthy Majuta (consultant coordinator, clinical nursing instructor and facilitator).

## Notes

**<u>Disclosures</u>** The authors declare no competing nor conflict of interests.

### Competing Interest Statement

The authors have declared no competing interest.

### Funding Statement

This study did not receive any funding

### Author Declarations

This project was determined to be exempt from Institutional Review Board review according to the IRB Secretary Dr. Fraja S. Chiwanga of the Muhimbili National Hospital Teaching, Research, and Consultancy Unit at the time of submission.

## References

1. United Nations. The 17 Sustainable Development Goals. UN. https://sdgs.un.org/goals

2. Frenk J, Chen L, Bhutta ZA, et al. Health professionals for a new century: transforming education to strengthen health systems in an interdependent world. The Lancet. 2010;376(9756):1923–1958. doi:10.1016/s0140-6736(10)61854-5

3. World Health Organization. Universal Health Coverage. WHO. https://www.who.int/health-topics/universal-health-coverage#tab=tab_1

4. Bvumbwe T, Mtshali N. Nursing education challenges and solutions in Sub Saharan Africa: an integrative review. BMC Nursing. 2018;17(3). doi:10.1186/s12912-018-0272-4

5. Mormina M, Pinder S. A conceptual framework for training of trainers (ToT) interventions in global health. Globalization and Health. 2018;14(1):100. doi:10.1186/s12992-018-0420-3

6. The Alliance. AGCT. https://www.agct.info/

7. Our Profile. The United Republic of Tanzania MUHIMBILI NATIONAL HOSPITAL (MNH). http://www.mnh.or.tz/index.php/our-profile

8. Peters A, Palomo R, Pittet D. The great nursing brain drain and its effects on patient safety. Antimicrobial Resistance & Infection Control. 2020;9(1):57. doi:10.1186/s13756-020-00719-4

9. United Nations. World population projected to reach 9.8 billion in 2050, and 11.2 billion in 2100. UN. Published 2017. https://www.un.org/en/desa/world-population-projected-reach-98-billion-2050-and-112-billion-2100

10. Ray ML, Wilson MM, Wandersman A, Meyers DC, Katz J. Using a Training-of-Trainers Approach and Proactive Technical Assistance to Bring Evidence Based Programs to Scale: An Operationalization of the Interactive Systems Framework’s Support System. American Journal of Community Psychology. 2012;50(3-4):415–427. doi:10.1007/s10464-012-9526-6

11. About Us | OER Africa. https://www.oerafrica.org. https://www.oerafrica.org/about-us

12. Toney-Butler TJ, Thayer JM. Nursing Process. StatPearls. StatPearls Publishing; 2021.

13. Tool: SBAR | Agency for Healthcare Research and Quality. https://www.ahrq.gov. https://www.ahrq.gov/teamstepps-program/curriculum/communication/tools/sbar.html

14. Mtakasimba S. WATAALAMU WA ICU MNH WAJENGEWA UWEZO. MNH Official Blog. Published August 29, 2019. http://www.mnh.or.tz/muhimbiliblog/story/wataalamu-wa-icu-mnh-%20wajengewa-uwezo-/

