## Appendix for "A Training-of-Trainers Program for Nurses in Tanzania: ICU Standards of Care, Documentation, and Communication"

Grace Kistner, RN, PDTN, MMHA, BSBA, BSN, CCRN, CSSLHPM & Shannon Macfarlan, BSN-RN, CPHN

**Supplemental Materials: Clinical Topics Knowledge Assessment Data Findings**

**Appendix 1: HEMODYNAMICS AND ECG INTERPRETATION**

The cardiovascular lecture began with a review of cardiac output and clinical nursing assessments used to determine if tissue perfusion is adequate. A normal sinus rhythm ECG strip with the associated measurements (PR, QRS, QT, R-R) was reviewed as a baseline patient assessment. An atrial fibrillation ECG was discussed at length, including the associated side effects (stable versus unstable) and treatments (medications and cardioversion). The participants were tested on the equation for cardiac output, components of stroke volume, non-invasive cardiac output assessments, ECG interpretation, hallmarks of and treatment for atrial fibrillation. The participants' average overall scores increased from 45% to 83% after the course (Appendix Figure 1a). A distribution of test scores (Appendix Figure 1b) showed 35% of the participants were able to successfully answer all of the questions before the course, whereas 46% could not answer any successfully. After the course, 70% answered all correctly.

**Appendix Figure 1a. Knowledge Assessment: Cardiac Assessment and Hemodynamics**

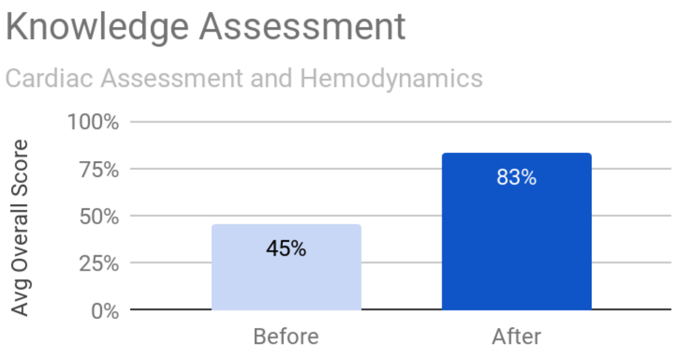


**Appendix Figure 1b. Distribution of Scores: Cardiac Assessment and Hemodynamics**

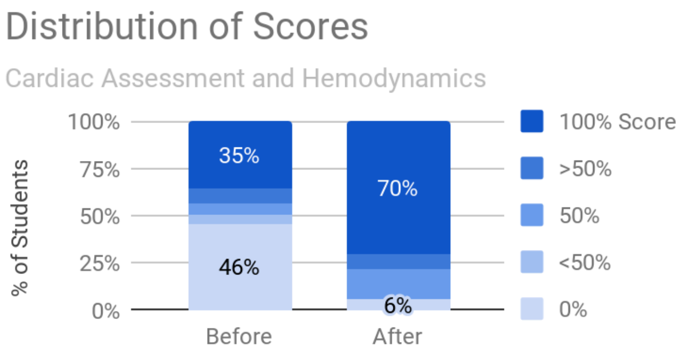

Grace Kistner, RN, PDTN, MMHA, BSBA, BSN, CCRN, CSSLHPM & Shannon Macfarlan, BSN-RN, CPHN

#### Appendix 2: PSAD MANAGEMENT AND STROKE

The neurology-focused lecture began with discussing the patient's baseline neurological exam in order to determine any subtle changes. The Glasgow Coma Scale and the patient's level of consciousness were reviewed, then the lecture advanced into motor and pupillary responses. The protective brainstem reflexes and their importance produced important discussions within the class. The definitions of ischemic versus hemorrhagic stroke and respective clinical presentations were reviewed. The acting FAST (Face, Arms, Speech and Time) for stroke assessment was reinforced and presented as an educational piece for patients and families. Pain, sedation, delirium and the implications with an ICU patient were discussed at length as well as the CAM-ICU assessment tool to determine if the patient is delirious. Providing adequate pain and sedation medications for the intubated ICU patient in order to maintain an appropriate RASS (Richmond Agitation Sedation Scale) was discussed as well as advocating patient's needs to the physicians. The participants' average overall scores increased from 73% to 92% after the course (Appendix Figure 2a). Before the course, 61% of the participants were able to correctly answer all test questions, compared to 89% (Appendix Figure 2b).

**Appendix Figure 2a. Knowledge Assessment:**
**Pain, Sedation, Agitation, and Delirium (PSAD) and Stroke**

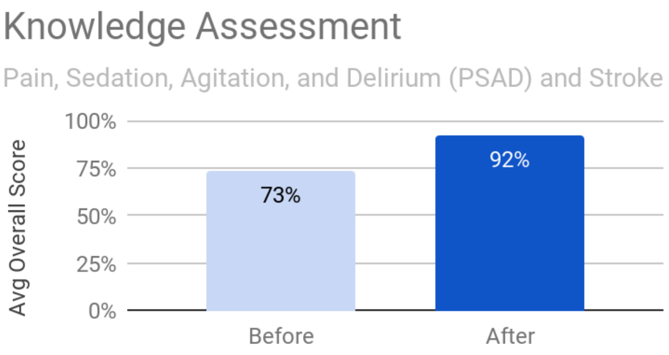


**Appendix Figure 2b. Distribution of Scores:**
**Pain, Sedation, Agitation, and Delirium (PSAD) and Stroke**

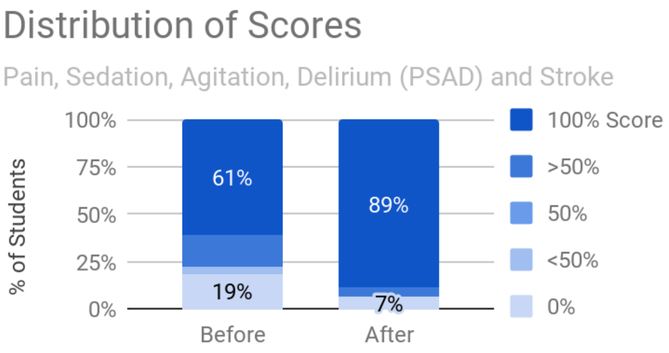

Grace Kistner, RN, PDTN, MMHA, BSBA, BSN, CCRN, CSSLHPM & Shannon Macfarlan, BSN-RN, CPHN

#### Appendix 3: VENTILATOR MANAGEMENT AND ARDS

The ventilator management and Acute Respiratory Distress Syndrome lecture began by discussing the pathophysiology of oxygen transport and delivery in the body. Clinical assessment of pulmonary mechanics, indications that necessitate intubation, and extubation criteria were discussed at length. Interpreting arterial blood gases (ABG) and the importance of monitoring peripheral capillary oxygen saturation (SpO2) were utilized to discuss causes of and treatment for hypoxia. Acute Respiratory Distress Syndrome (ARDS) and corresponding ventilator settings were highlighted to reinforce the key of customizing nursing care plans. The participants' average overall scores increased from 78% to 82% after the course (Appendix Figure 3a). Baseline ventilator comprehension was indicated by 63% of the participants being able to successfully answer all of the test questions before the course; and the content review increased that number to 73% after the course (Appendix Figure 3b).

**Appendix Figure 3a. Knowledge Assessment:**
**Airway and Ventilator Management**

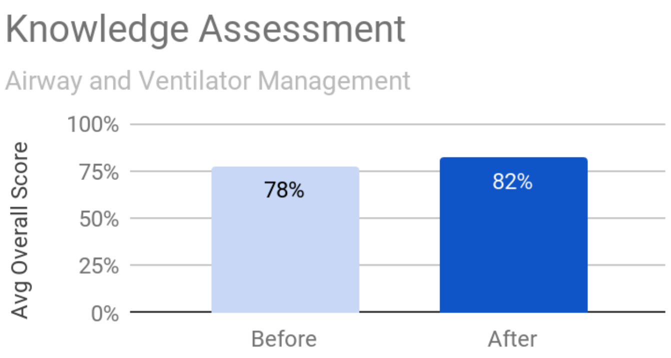


**Appendix Figure 3b. Distribution of Scores:**
**Airway and Ventilator Management**

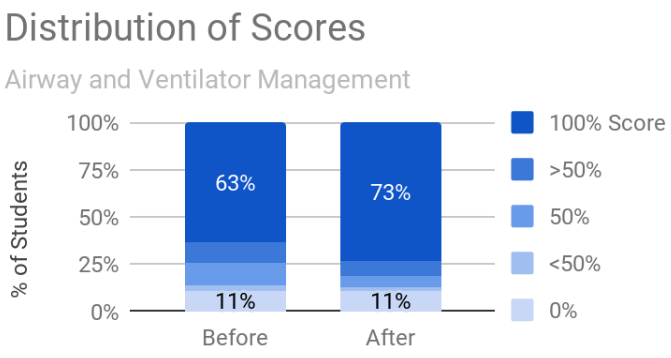

#### Appendix 4: METABOLIC DISORDERS AND NUTRITION

The principles of preventing acute kidney injury (AKI) as well as the associated assessments to determine kidney injury were discussed in the metabolic disorders lecture. Risk factors for and types of kidney injury (pre-renal, renal, and post-renal) were reviewed, including the treatments available. In addition, the importance of starting early, balanced nutrition was emphasized as well as contraindications for nutrition therapy. Finally, routine blood sugar maintenance and the implications of hypo- and hyperglycemia in a critically ill patient was discussed, focusing on diabetic ketoacidosis management. The participants' average overall scores increased from 58% to 93% after the course (Appendix Figure 4a). A great deal of learning was indicated by an increase from 56% to 93% of the participants being able to successfully answer all test questions before and after the course (Appendix Figure 4b).

**Appendix Figure 4a. Knowledge Assessment: Metabolic Disorders and Nutrition**

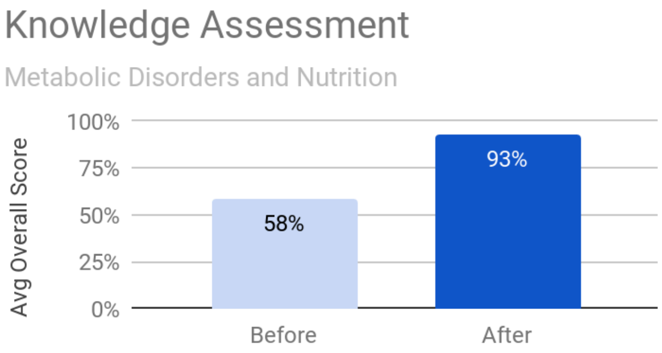


**Appendix Figure 4b. Distribution of Scores: Metabolic Disorders and Nutrition**

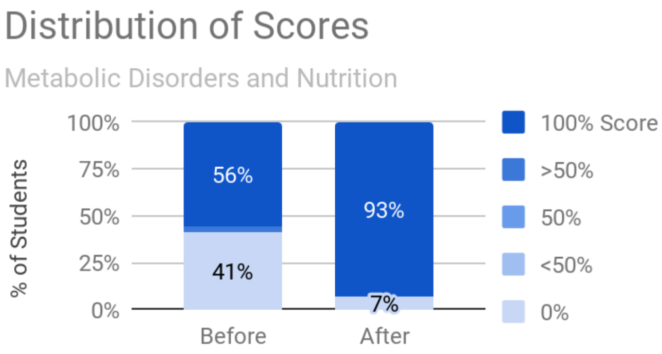

Grace Kistner, RN, PDTN, MMHA, BSBA, BSN, CCRN, CSSLHPM & Shannon Macfarlan, BSN-RN, CPHN

### Appendix 5: ICU BUNDLES AND STANDARDS OF CARE AND DOCUMENTATION

The ICU bundles were discussed as standards of care for the ICU patient, such as Deep Vein Thrombosis and Ventilator Associated Pneumonia prophylaxis. Standard universal precautions, which included differentiating between single versus multiple use items, were emphasized as well as infection control precautions and regular assessments of invasive devices. Discussions reviewed providing close monitoring of surgical sites to determine the wound's healing and communicating abnormalities to the physician. The detailed assessment, staging and documentation of pressure injuries as well as nursing interventions for prevention (such as turning every two hours and elevating bony prominences from the bed) was discussed in detail. The participants' average overall scores increased from 66% to 85% after the course (Appendix Figure 5a). This review in basics of standard care improved knowledge, as evidenced by 41% to 67% of the participants being able to successfully answer all test questions before versus after the course (Appendix Figure 5b).

**Appendix Figure 5a. Knowledge Assessment:**
**ICU Bundles and Standards of Care**

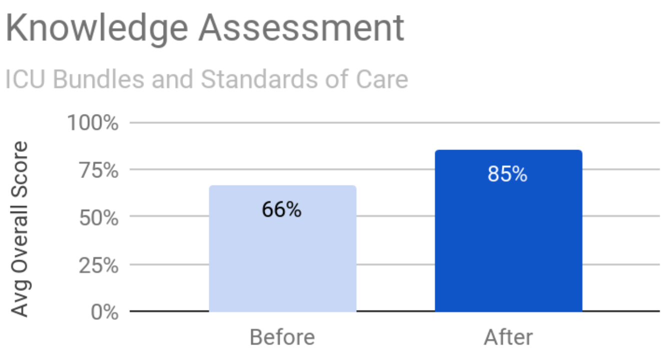


**Appendix Figure 5b. Distribution of Scores:**
**ICU Bundles and Standards of Care**

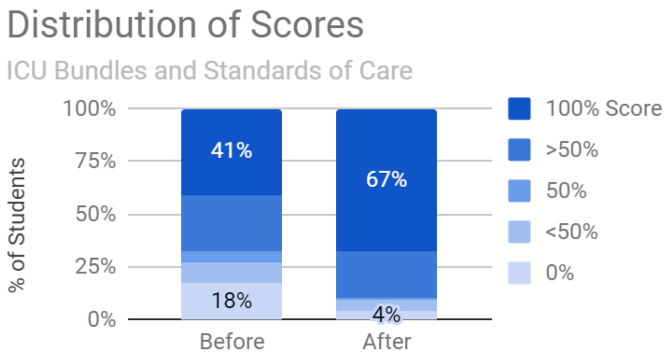

### Appendix 6: MODS, SEPSIS, AND END OF LIFE

The Multiple Organ Dysfunction Syndrome, sepsis, palliative care, and end-of-life care lecture emphasized recognition and early implementation of sepsis treatment, as well as the implications that can occur in multiple body systems. Early sepsis management focused on the one-hour bundle of source identification and control, empiric broad-spectrum antibiotic therapy and fluid resuscitation. The evidence of Multiple Organ Dysfunction Syndrome by body system was reviewed and inclusion criteria provided. Palliative care was carefully defined and the ethical issues ICU nurses face at the bedside was discussed at length. Legal components involving code status directed therapies and cultural considerations regarding family involvement in decision making was compared between US and Tanzanian practices. A supportive and open discussion highlighted the moral distress that this can cause for direct patient care providers. The collaboration in the care for patients with grave prognosis with the medical team was emphasized. The participants' average overall scores increased from 67% to 94% after the course (Appendix Figure 6a). Results of the distribution of scores for this course topic showed the most significant amount of learning gained with a 52% increase in the participants being able to successfully answer all questions (Appendix Figure 6b).

**Appendix Figure 6a. Knowledge Assessment: Sepsis and Palliative Care**

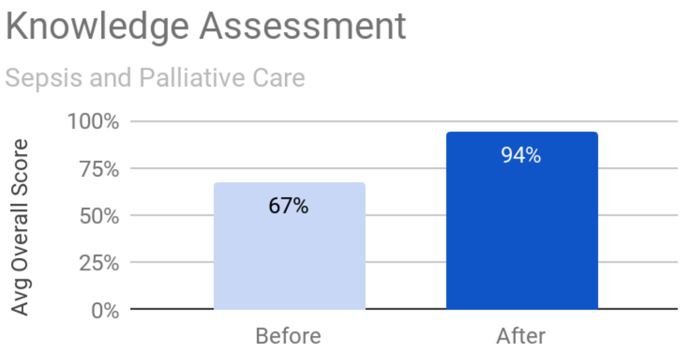


**Appendix Figure 6b. Distribution of Scores: Sepsis and Palliative Care**

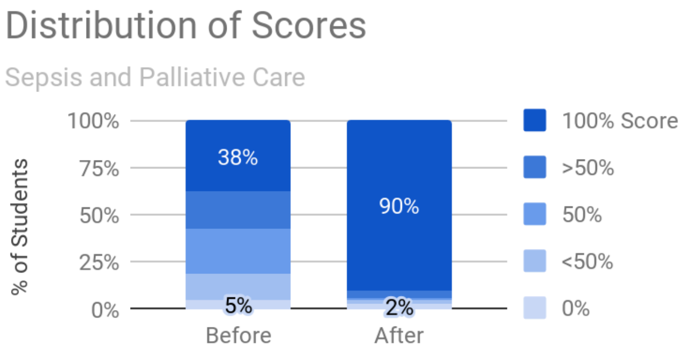
